# Insights from the second season of collaborative influenza forecasting in Italy with updated targets incorporating virological information

**DOI:** 10.64898/2026.03.04.26347601

**Authors:** Stefania Fiandrino, Tommaso Bertola, Valeria D’Andrea, Manlio De Domenico, Elisa Viola, Lorenzo Zino, Mattia Mazzoli, Alessandro Rizzo, Yuhan Li, Nicola Perra, Marika Sartore, Razieh Masoumi, Chiara Poletto, Alberto Mateo Urdiales, Antonino Bella, Corrado Gioannini, Paolo Milano, Daniela Paolotti, Marco Quaggiotto, Luca Rossi, Ivan Vismara, Alessandro Vespignani, Nicolò Gozzi

## Abstract

We present results from the second season of Influcast, a multi-model collaborative forecasting hub focused on influenza in Italy. During the 2024/25 winter season, Influcast collected one- to four-week-ahead probabilistic forecasts of influenza-like illness (ILI) incidence alongside influenza A and B *ILI+* incidence signals. New ILI+ targets were constructed integrating syndromic surveillance data with virological detections collected weekly by the Italian National Institute of Health. Forecasts were submitted by six independent models (including compartmental, metapopulation, and statistical approaches) and combined into an ensemble. Ensemble forecasts for ILI+ consistently outperformed both the baseline (a naive persistence model) and most individual models in terms of Weighted Interval Score (WIS), Absolute Error (AE), and prediction coverage. Importantly, ensemble ILI+ forecasts achieved significantly lower WIS and AE ratios (i.e., ratio between the ensemble and the baseline models) and improved calibration compared to ILI forecasts. Our findings support the integration of virological surveillance data in forecasting target definition to improve the reliability of epidemic forecasts and strengthen their utility for situational awareness, communication, and targeted intervention.

## 1 Introduction

Influenza-like illness (ILI) is a syndromic indicator routinely collected by public health authorities to monitor the burden of seasonal respiratory pathogens [1–3]. Since ILI incidence provides timely signals of community-level transmission, it is commonly used as target in forecasting models that aim to anticipate the dynamics of seasonal influenza epidemics [4–6]. However, ILI is defined through the co-occurrence of symptoms, such as fever and cough, rather than confirmed pathogen detection, and thus does not uniquely identify influenza cases [7–10]. In practice, ILI reflects the aggregate incidence of multiple, overlapping respiratory infections with similar clinical presentations. The specificity of ILI for detecting influenza has further declined since the emergence of SARS-CoV-2, which has added to the mix of co-circulating pathogens contributing to ILI counts [11–13].

From a forecasting perspective, using targets that involve multiple underlying epidemiological processes may obscure distinct dynamics, leading to noisier and less predictable signals. Consequently, models trained on ILI data may capture composite trends rather than pathogen-specific trajectories. This limits their ability to disentangle changes in influenza transmission from those driven by other co-circulating viruses, and ultimately reduces forecasting performance [14, 15].

In parallel, the expansion of virological surveillance systems providing quantitative estimates of pathogen-specific circulation has created new opportunities to move beyond purely syndromic indicators. These advancements enable the development of more informative forecasting targets that help models disentangle influenza activity from other respiratory infections [16–18].

Recent studies demonstrate that integrating virological data to refine forecasting targets enhances influenza forecast accuracy and informativeness [14, 19–21]. In the United States, forecasts stratified by influenza type and subtype were found to outperform aggregated models, yielding more accurate predictions both at the subtype and total incidence levels [19]. Similarly, mechanistic models trained on type-specific data demonstrated superior accuracy compared to those using total specimen data, particularly at longer lead times and finer spatial resolutions [20]. In parallel, approaches integrating multiple data streams have also proven highly effective. For instance, a model combined limited surveillance data with complementary signals such as the proportion of outpatient visits attributable to influenza and rates of laboratory-confirmed influenza hospitalizations, achieving top performance in the US CDC’s influenza prediction challenge in season 2023/24 [21]. Finally, further work showed that decomposing the ILI signal into viral components and forecasting each pathogen individually before aggregating the predictions can substantially improve both short-term and seasonal forecast accuracy [14].

In this study, we aim to evaluate whether incorporating virological data to refine forecasting targets can enhance predictive accuracy in the context of Influcast, a collaborative influenza forecasting hub operating in Italy [5]. Collaborative forecasting hubs are widely employed by researchers and public health institutions worldwide to generate short-term forecasts of infectious disease epidemics [22]. Their effectiveness largely emerges from the use of ensemble approaches, which combine multiple independent model submissions into a single aggregated prediction [23]. Ensemble forecasts have consistently been found to outperform individual model forecasts, showing greater robustness and accuracy in previous forecasting efforts [4–6, 24, 25]. However, limited research has examined how refining forecasting targets using virological information affects performance within multi-model collaborative hubs. Our work addresses this gap by systematically evaluating whether virologically informed targets yield measurable performance improvements in both ensemble and individual model forecasts.

During the 2024/25 winter season, we constructed weekly ILI+ signal, defined as the product of influenza-like illness incidence and the proportion of laboratory-confirmed influenza cases [16], for influenza type A and type B at the national level in Italy by integrating syndromic and virological surveillance data collected by the Italian National Institute of Health. Each week, independent modeling teams submitted probabilistic forecasts for ILI+ incidence signals over a four-week horizon, alongside analogous forecasts for ILI incidence. Individual forecasts were then combined into an ensemble and evaluated using the Weighted Interval Score, the Absolute Error and the prediction coverage as evaluation metrics.

Our results show that (i) the ensemble consistently ranked among the top-performing models for the new ILI+ incidence targets, outperforming both the baseline and most individual models across multiple scoring metrics, and (ii) integrating virological data into target definitions improved predictive accuracy across all forecast horizons and evaluation metrics for the ensemble and for most individual models.

Overall, we assess the added value of integrating virological and syndromic surveillance data to improve performance of short-term ensemble influenza forecasts and increase their operational relevance for real-time public health assessment and response.

## 2 Results

### 2.1 Forecasting challenge and contributing models

During the 2024/25 winter season, Influcast collected forecasts for three national-level targets in Italy: standard ILI incidence, defined as the number of reported ILI cases per 1, 000 patients, and two additional *ILI+* targets obtained by combining syndromic surveillance with virological detections. Specifically, *ILI+(Flu A)* represents the weekly incidence of ILI cases per 1,000 patients attributable to influenza virus type A, while *ILI+(Flu B)* refers to the corresponding incidence attributable to influenza virus type B. Further details on the forecasting targets definition are provided in Section 4.2.

Six independent models contributed one-to four-week-ahead probabilistic forecasts for the three targets over 18 forecasting rounds from week 48 of 2024 to week 13 of 2025. Submitted models included five mechanistic models, of which one using a metapopulation approach, and one statistical model. Additional details on participating models are provided in the Supplementary Information (Section S1). Forecasts were combined into an ensemble and compared against a naive baseline model (i.e., a persistence model used as a neutral benchmark), considering the Weighted Interval Score (WIS), the Absolute Error of the median (AE), and the prediction coverage. Further details on ensemble/baseline forecasts calculation and forecast evaluation are provided in Sections 4.3 and 4.4.

We note that 6 additional models submitted forecasts for the ILI incidence target only. Due to the earlier start of syndromic surveillance relative to virological surveillance, forecasts for ILI incidence started in week 45 of 2024, resulting in three additional rounds compared to the ILI+ targets. To ensure a fair comparison between ILI and ILI+ forecast performance, in the following analyses, we build ensembles considering only models that submitted all three targets. Additionally, we limit comparison to the rounds common to all targets. In the Supplementary Information, we provide extensive sensitivity analysis on model and forecasting round inclusion, showing minimal impact on the main findings.

### 2.2 Individual and ensemble models performance on ILI+ targets

First, we evaluate the performance of ensemble and individual model forecasts on new ILI+ targets. We initially focus on aggregated performance in terms of median relative WIS, relative AE, 50%, and 90% prediction coverage. The relative WIS is computed using the methodology described in Sec. 4.4, by aggregating forecasts from all rounds and horizons, comparing each model to the other models submitting forecasts in the same round, and finally considering the ratio between the WIS of a model and that of the baseline. It follows that relative WIS below (above) 1 indicates better (worse) performance compared to the baseline. An analogous quantity can be computed considering the AE of the median as evaluation metric. Prediction coverage is instead defined as the fraction of observed values falling within a specified prediction interval (i.e., 90%). The prediction coverage is computed by pooling together forecasts from all rounds and horizons. Further details on forecast evaluation metrics are provided in Section 4.4.

Table 1 reports aggregated performance of different models in the 2024/25 season for the ILI+(Flu A) target. When considering the median relative WIS, the *Ensemble* achieves the second lowest value (0.38), following closely *Mechanistic-1* (0.35) and followed by *Mechanistic-3* (0.43). Next are *Mechanistic-3* (0.54), *Mechanistic-5* (0.60), *Mechanistic-4* (0.80), and *Statistical-1* (0.91), all outperforming the *Baseline* model.

**Table 1:** Aggregated forecasting performance of different models on ILI+(Flu A), season 2024/25. Performance of different models in terms of relative WIS, relative AE of the median, 50%, and 90% coverage. For each metric, the score of the model achieving the best performance is highlighted in bold, and the ranking of the top three models is indicated in parentheses.

| Model | Relative WIS | Relative AE | Coverage (50%) | Coverage (90%) |
| --- | --- | --- | --- | --- |
| *Mechanistic-1* | **0.35** (1<sup>st</sup>) | **0.39** (1<sup>st</sup>) | 0.28 | **0.60** (1<sup>st</sup>) |
| *Ensemble* | 0.38 (2<sup>nd</sup>) | 0.40 (2<sup>nd</sup>) | 0.40 (2<sup>nd</sup>) | **0.60** (1<sup>st</sup>) |
| *Mechanistic-3* | 0.43 (3<sup>rd</sup>) | 0.41 (3<sup>rd</sup>) | 0.29 | 0.58 (2<sup>nd</sup>) |
| *Mechanistic-2* | 0.54 | 0.54 | **0.42** (1<sup>st</sup>) | 0.53 (3<sup>rd</sup>) |
| *Mechanistic-5* | 0.60 | 0.55 | 0.17 | 0.47 |
| *Mechanistic-4* | 0.80 | 0.77 | 0.07 | 0.24 |
| *Statistical-1* | 0.91 | 0.88 | 0.28 | 0.49 |
| *Baseline* | 1.00 | 1.00 | 0.29 | 0.58 (2<sup>nd</sup>) |

Considering the relative AE, *Mechanistic-1* is the top-performing model (0.39), followed closely by the *Ensemble* (0.40), while *Mechanistic-3* ranks third (0.41). The remaining models, *Mechanistic-2* (0.54), *Mechanistic-5* (0.55), *Mechanistic-4* (0.77), and *Statistical-1* (0.88), also show values below 1, indicating improved performance compared to the *Baseline*.

When considering prediction coverage, the *Ensemble* achieves the closest value to nominal coverages for the 90% prediction interval (0.60), and the second closest value for 50% prediction interval (0.40). *Mechanistic-1* matches the *Ensemble* in 90% coverage (0.60), followed by the *Baseline* and *Mechanistic-3* (0.58), and *Mechanistic-2* (0.53). For the 50% coverage, the *Ensemble* is followed by *Mechanistic-2* (0.40) and the *Baseline* (0.34).

Table 2 shows similar findings for the ILI+(Flu B) forecasting target. In this case, *Mechanistic-3* is the top-performing model according to all evaluation metrics considered, except for the 50% coverage. The *Ensemble* ranks second in terms of relative WIS, relative AE, and 90% coverage, and first for 50% coverage. Of the remaining models, *Mechanistic-1* shows competitive performance, ranking third for the relative WIS and relative AE, and for the two prediction coverage levels considered. In contrast, *Mechanistic-5* struggles to outperform the baseline across the different evaluation metrics.

**Table 2:** Aggregated forecasting performance of different models on ILI+(Flu B), season 2024/25. Performance of different models in terms of relative WIS, relative AE of the median, 50%, and 90% coverage. For each metric, the score of the model achieving the best performance is highlighted in bold, and the ranking of the top three models is indicated in parentheses.

| Model | Relative WIS | Relative AE | Coverage (50%) | Coverage (90%) |
| --- | --- | --- | --- | --- |
| *Mechanistic-3* | **0.36** (1<sup>st</sup>) | **0.44** (1<sup>st</sup>) | 0.32 | **0.74** (1<sup>st</sup>) |
| *Ensemble* | 0.41 (2<sup>nd</sup>) | 0.45 (2<sup>nd</sup>) | **0.36** (1<sup>st</sup>) | 0.64 (2<sup>nd</sup>) |
| *Mechanistic-1* | 0.52 (3<sup>rd</sup>) | 0.54 (3<sup>rd</sup>) | 0.33 (3<sup>rd</sup>) | 0.63 (3<sup>rd</sup>) |
| *Mechanistic-2* | 0.70 | 0.67 | 0.22 | 0.43 |
| *Mechanistic-4* | 0.84 | 0.88 | 0.14 | 0.39 |
| *Statistical-1* | 0.84 | 0.74 | 0.35 (2<sup>nd</sup>) | 0.44 |
| *Baseline* | 1.00 | 1.00 | 0.18 | 0.44 |
| *Mechanistic-5* | 1.01 | 0.91 | 0.06 | 0.14 |

Figure 1 shows the distribution of standardized WIS ranks across all forecasting horizons and rounds for each model for ILI+(Flu A) and ILI+(Flu B) targets. The standardized rank is defined such that the best-performing model for a given horizon and round receives a score of 1 and the worst-performing model a score of 0 (see Section 4.4). For ILI+(Flu A), the *Ensemble* achieves the highest median standardized rank, followed by *Mechanistic-1* and *Mechanistic-3*, which share the same median value. For ILI+(Flu B), the *Ensemble* ranks second in terms of median standardized WIS rank, following *Mechanistic-3*, and being followed by *Mechanistic-1* and *Mechanistic-2*. These findings confirm the results from Tables 1 and 2. The *Ensemble* not only achieves highest or close-to-highest median standardized ranks but also most frequently appears in the top half of the ranking distribution, exceeding 80% of rounds for ILI+(Flu A) and 90% for ILI+(Flu B). In other words, a greater fraction of the *Ensemble*’s WIS rank distribution lies above 0.5, suggesting superior stability in its performance across rounds compared to other top-performing models. The corresponding rank performance analysis based on the absolute error of the median is provided in Figure S1 of the Supplementary Information. When evaluated using this metric, the Ensemble model demonstrates stable performance, ranking within the top half of the distribution in 78% of rounds for ILI+(Flu A) and 88% for ILI+(Flu B).

**Figure 1.**
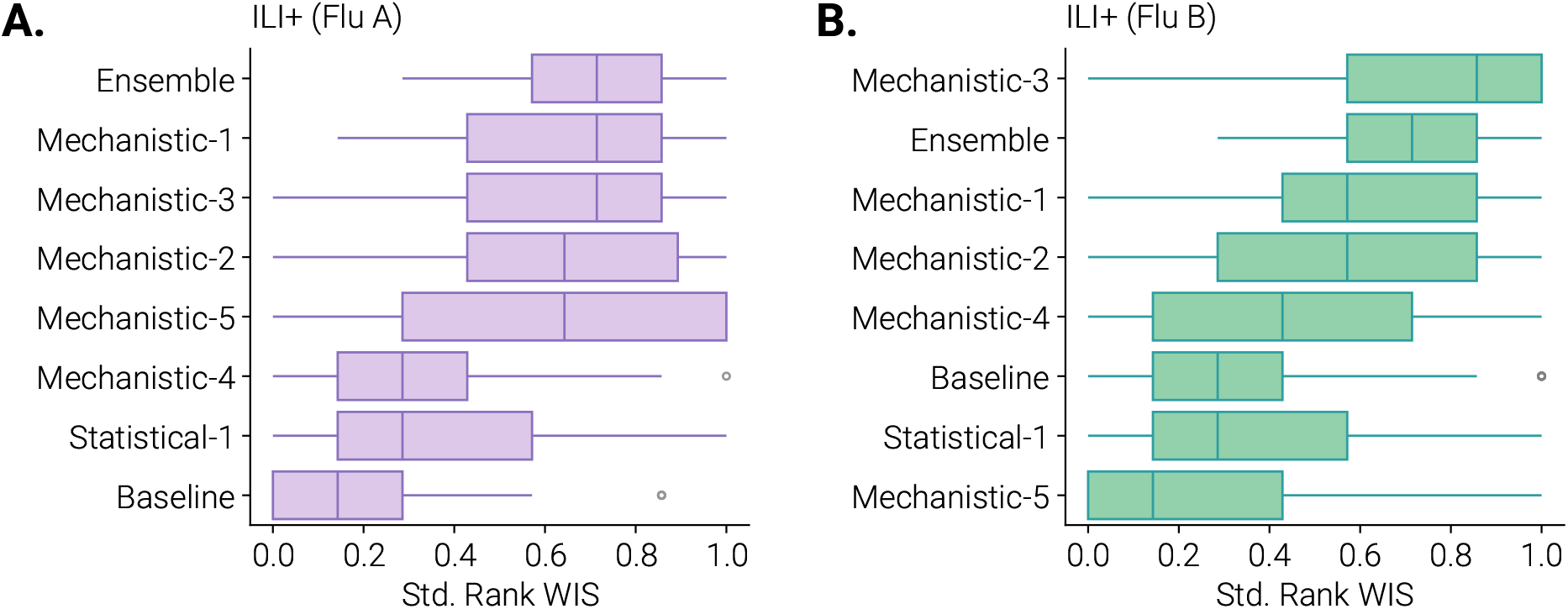
Standardized WIS rank of 2024/2025 Influcast forecasting models for ILI+(Flu A) and ILI+(Flu B). The plot shows boxplots of standardized WIS ranks of different models computed by combining different horizons and forecasting rounds for ILI+(Flu A) (panel A) and ILI+(Flu B) (panel B). The boxplot boundaries represent the interquartile (IQR) range spanning between the first (Q1) and third (Q3) quartile, while the line inside each box indicates the median. The whiskers extend to the furthest data point within 1.5 times the IQR from Q1 and Q3. Outliers are shown as individual points.

### 2.3 Comparison between ILI and ILI+ ensemble forecasts performance

To assess whether incorporating virological information in forecasting target definition improves predictive accuracy, we compared the performance of ensemble and individual models forecasts of ILI and ILI+ incidence targets.

Figure 2A shows distributions, aggregated and separated by forecast horizon, of the WIS ratio between the Ensemble and the Baseline for ILI, ILI+(Flu A), ILI+(Flu B). When considering aggregated performance across all horizons (left panel), the median ratio of the WIS between the Ensemble and Baseline models on the ILI target is consistently higher than that of the ILI+(Flu A) and ILI+(Flu B) ensembles (0.82 compared to 0.36 and 0.47, respectively). Similar patterns are observed when performance is analyzed by forecast horizon: the median ratio of the WIS between the Ensemble and Baseline models of the ILI ensemble exceeds that of both ILI+ ensembles across all four horizons, with differences more pronounced for ILI+(Flu A). As the forecast horizon increases, differences in the median ratio gradually diminish. For example, at horizon 1 the median ratio of the ILI ensemble is 1.04, compared to 0.41 and 0.73 for the ILI+(Flu A) and ILI+(Flu B) ensembles, respectively, corresponding to absolute differences of 0.63 and 0.31. At horizon 4, these differences reduce to 0.24 and 0.13. To assess the statistical significance of these differences, we applied the Wilcoxon signed-rank test to the full distribution of the WIS ratios, performing two pairwise comparisons: ILI vs ILI+(Flu A) and ILI vs ILI+(Flu B). When aggregating across all horizons, the differences are significant in both cases at the 1% significance level. When disaggregated by horizon, for ILI+(Flu A), the difference remains significant at the 1% level for horizons 1 and 2, at the 5% level for horizon 3, and at the 10% level for horizon 4. For ILI+(Flu B), the difference is significant at the 10% level for all horizons except horizon 1, where it is not significant. The reduced significance at the horizon-specific level also reflects smaller sample sizes when disaggregating by horizon.

**Figure 2.**
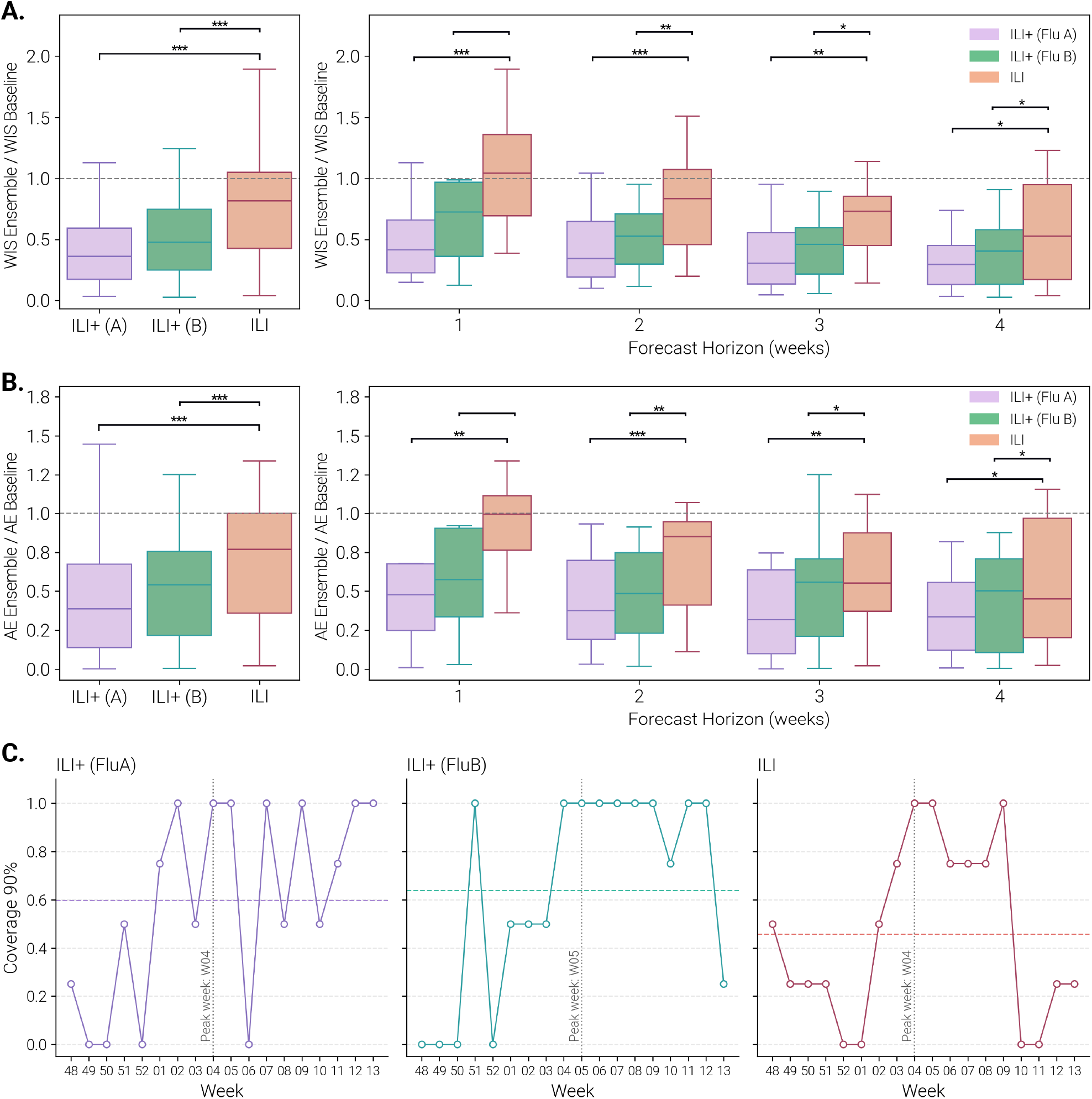
Influcast Ensemble forecasting performance on ILI and ILI+. WIS ratio between the ensemble and baseline models **(A)** and AE ratio between the ensemble and baseline models **(B)** computed aggregating across forecasting rounds and horizons (left panels) and disaggregated by horizons (right panels). Boxplots show the distribution of WIS/AE values. The boxplot boundaries represent the interquartile range (IQR) between the first quartile (Q1) and third quartile (Q3), while the line inside each box indicates the median. The whiskers extend to the furthest data point within 1.5 times the IQR from Q1 and Q3. In all the panels, we report the statistical significance of the Wilcoxon signed-rank test comparing ILI versus ILI+ (Flu A) and ILI+ (Flu B) as follows: ^***^: *p*_*val*_ *<* 0.01, ^**^: 0.01 *< p*_*val*_ *<* 0.05, ^*^: 0.05 *< p*_*val*_ *<* 0.1, and blank otherwise. **(C)** 90% coverage values in different rounds for ILI, ILI+(Flu A), and ILI+(Flu B). Dashed horizontal lines represent the average value across forecasting rounds, while the grey vertical dashed line indicates the week of the peak of each target.

Figure 2B reports analogous results for AE ratio between the Ensemble and the Baseline, showing consistent patterns. At the horizon-aggregated level (left panel), the ILI ensemble performs worse than both ILI+ ensembles in terms of the median ratio of the AE between the Ensemble and Baseline models, with differences significant at the 1% level. When analyzed by horizon, the ILI ensemble shows higher median ratio of the AE between the Ensemble and Baseline models than both ILI+ ensembles across all horizons except horizon 4 for ILI+(Flu B). As in the case of WIS, the performance gains of ILI+ ensembles relative to ILI diminish with increasing horizon, particularly for ILI+(Flu B). Horizon-level Wilcoxon signed-rank tests show results consistent with the WIS analysis: ILI+(Flu A) shows significant differences at the 1% level for horizons 2, at the 5% level for horizon 1 and 3, and at the 10% level for horizon 4, while differences for ILI+(Flu B) are significant at the 5% level for horizon 2, at the 10% level for horizons 3 and 4, and not significant for horizon 1.

Figure 2C shows the 90% prediction coverage values in different forecasting rounds, separately for the three targets. For each round, we computed the average coverage across the four horizons in that round. The dashed horizontal lines represent the average coverage across all rounds, with mean values of 0.46 for ILI, 0.60 for ILI+(Flu A), and 0.64 for ILI+(Flu B), indicating, on average, improved calibration of the ensemble for the ILI+ targets. Examining temporal trends, we observed that ensemble forecasts were systematically overconfident during the early forecasting rounds leading up to the epidemic peak, with the 90% prediction intervals bounding observed incidence less frequently than expected (i.e., prediction coverage falling below nominal levels). This overconfidence was most pronounced for the ILI incidence target. After the peak, coverage generally improved for all targets, approaching nominal levels, before declining slightly toward the end of the season, particularly for ILI.

The comparison of individual model performance across ILI and ILI+ targets is presented in Section S3 of the Supplementary Information. Overall, most individual models benefit from incorporating virological information in terms of improved predictive performance. All six individual models show higher median ratio of the WIS and AE between the ensemble and baseline models for ILI than for ILI+(Flu A), with such differences being statistically significant in three cases for both metrics. For ILI+(Flu B), three models show higher median ratio of the WIS and AE between the ensemble and baseline models for ILI, with statistical significance in two cases.

## 3 Discussion

This study investigated the impact of incorporating virological information in forecasting target definition on the predictive performance of ensemble in an operational forecasting context. We explored to what extent forecasting performance gains associated with a cleaner by design ILI+ signal are consistently observed across an entire influenza season, multiple independent individual models, and their ensemble within the same forecasting challenge.

During the 2024/25 winter season, Influcast supported three forecasting targets: the standard ILI incidence, commonly used to predict the dynamics of seasonal influenza epidemics, and two additional ILI+ incidence indicators that integrate syndromic and virological surveillance information, for both influenza type A and type B. Six different models submitted four-week-ahead probabilistic forecasts for the three targets over 18 consecutive rounds, from November 2024 to the end of March 2025. When evaluated on novel ILI+ targets, the ensemble ranked among the top-performing models across different evaluation metrics, consistently outperforming the baseline and most individual models. These findings confirm results from the 2023/24 Influcast season [5] and reinforce the relevance and effectiveness of multi-model ensembling efforts also in the case of refined epidemiological targets fusing syndromic and virological surveillance.

The comparison between ILI and ILI+ ensemble performance suggests that integrating virological information in target definition contributes to improved predictive accuracy. This finding holds across different evaluation metrics, although the improvement is larger for ILI+(Flu A), tends to diminish as the forecast horizon increases, and is not consistently observed across all individual models. Smaller sample sizes at each horizon likely contribute to the reduced significance observed in horizon-specific analyses. The weaker performance observed for ILI+(Flu B) may reflect the lower and delayed circulation of influenza B compared to influenza A during the study period.

The present study comes with limitations. First, our findings are based on a single season and geographical scale. Future work could extend this assessment across multiple seasons to account for inter-seasonal variability and mitigate potential biases specific to a single season. With increasing availability of virological information, we may also be able to construct ILI+ targets at the sub-national level and extend the comparison also at more granular spatial scales. Second, the delayed release of virological surveillance data (starting on week 48, 2024) compared to syndromic data (week 45, 2024) resulted in three early forecasting rounds without ILI+ forecasts. While this represents a minor portion of all evaluated rounds (3/21 rounds), it may slightly affect the comparative performance assessment. Since no retrospective virological data were available to reconstruct these early rounds, we conduct the evaluation of the common rounds where both ILI and ILI+ forecasts were available, ensuring consistency in the comparison. Third, the number of models contributing forecasts for ILI+ targets was limited compared to the number of models submitting for ILI incidence. Nevertheless, the number of included models in this study meets the minimum requirement to construct an effective ensemble, as shown by previous research [26]. Lower participation for the new targets was expected, as the 2024/25 season was the first to include them, which required additional effort from participating modeling teams. As a result, some teams needed more time to prepare their submissions and missed the few initial rounds for the ILI+ targets. To address this, we invited retrospective integration of the missing rounds. Although these retrospective forecasts do not fully replicate real-time operational conditions, they were produced using only data available at the time of forecast submission (i.e., non-backfilled data), thereby reproducing the real-time forecasting setup as closely as possible. Beyond the number of participating models, the heterogeneity of approaches was also limited, with most models being mechanistic. Future work could introduce incentives for submitting alternative modeling approaches to assess whether greater methodological diversity may further enhance forecasting performance on new refined targets. Finally, the ensemble technique adopted in this study is intentionally simple and exploring alternative ensembling strategies could further improve predictive performance on new targets.

In conclusion, our findings support the inclusion of virological information in forecasting target definition to achieve improved short-term predictive performance. Beyond accuracy, these refined targets also have the potential to better support public health decision-making and risk communication. By being more pathogen-specific, they provide clearer insights into the dynamics of co-circulating pathogens during the winter season, enabling more precise communication and targeted interventions. While the ILI+ indicators for influenza A and B were developed as a proof of concept during the 2024/25 season, the same framework can be extended to other respiratory pathogens that contribute to the ILI burden. ILI surveillance captures infections caused by multiple respiratory pathogens whose circulation may differ in timing, intensity, and predictability. Data from the Italian RespivirNet surveillance system indicate that, in addition to influenza, pathogens such as RSV, rhinovirus, seasonal coronaviruses, SARS-CoV-2, adenovirus, parainfluenza virus, human metapneumovirus, and bocavirus contribute to the observed ILI signal. The co-circulation of these pathogens likely introduces additional variability into the ILI time series that is not explicitly captured by influenza-focused models, and may contribute to the lower forecast performance observed for ILI relative to ILI+. This finding highlights the potential value of integrating a broader set of virological signals as data availability improves. Future work could therefore move toward a comprehensive multi-pathogen forecasting approach, explicitly incorporating additional virological indicators alongside influenza. Moreover, alternative syndromic indicators, such as acute respiratory infection (ARI), which differ from ILI in sensitivity and specificity for non-influenza pathogens, could be also incorporated to further refine target definitions. An additional promising direction is the reconstruction of aggregate ILI forecasts from pathogen-specific predictions, allowing modeling teams to focus on mechanistically coherent targets while deriving the overall ILI signal through a centralized post-processing step.

Overall, these findings highlight the value of continuously refining epidemic forecasting targets to improve predictive accuracy and provide clearer, more actionable guidance for public health responses in the context of co-circulating respiratory viruses.

## 4 Materials and methods

### 4.1 Surveillance data and Influcast infrastructure

We used data on ILI incidence and virological detections collected through the national integrated surveillance system *RespiVirNet* [27]. *RespiVirNet* is coordinated by the Italian National Institute of Health with the support of the Italian Ministry of Health.

*RespiVirNet* integrates syndromic and virological surveillance of respiratory viruses, including influenza, through a national network of general practitioners, pediatricians, and regional reference laboratories. These actors report, on each week during the winter season, new ILI cases and laboratory-confirmed detections of a range of pathogens across all Italian regions and autonomous provinces.

Syndromic surveillance in season 2024/25 started on week 42, 2024, with the first bulletin published on week 45 of 2024, while virological surveillance started on week 46 with data being released from week 48 onward. The surveillance period ended in week 17 of 2025. Surveillance data were typically released every Friday, with new reported data referring to the preceding week. The Influcast platform was updated on the following Wednesday, allowing participating teams time to process the newly available data and submit forecasts for the new round. As a result, forecasts published on Influcast each Wednesday included forecasts for the previous week (for which consolidated surveillance data was not yet available), the current week, and the two subsequent weeks. We evaluate 18 forecasting rounds (2024-48 to 2025-13), excluding those missing full 4-week-ahead horizon. We refer to Ref. [5] for further details on the Influcast platform, including forecast submission format, validation procedures, and general description of the forecasting challenge.

### 4.2 Forecasting targets description

During the 2024/25 season, Influcast collected short-term forecasts of three different targets.

#### ILI incidence

The ILI incidence forecasting target is defined as the number of new weekly ILI cases reported by the network of sentinel general practitioners and pediatricians per 1,000 patients in Italy (including all regions and autonomous provinces). This indicator serves as the primary target of the Influcast forecasting challenge and it is directly communicated by the Italian National Institute of Health through the *RespiVirNet* weekly bulletin [27].

#### ILI+ incidence

The second set of forecasting targets, denoted as ILI+, represents the weekly incidence of ILI cases attributable to influenza viruses (type A and B). These quantities refine the ILI incidence indicator by integrating virological surveillance data [28]. More specifically, we consider information on the number of samples (collected through virological surveillance) that test positive for influenza viruses at the national level. This information is also provided, each week, in the *RespiVirNet* bulletin. For influenza virus type A, the corresponding ILI+ incidence indicator is computed as:

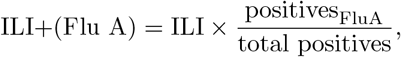

where ILI is the reported weekly ILI incidence, positives_FluA_ is the number of samples testing positive for influenza virus type A, and total positives is the total number of samples testing positive to monitored respiratory pathogens in the given week. Similarly, we can compute ILI+(Flu B) by considering the number of positive samples to the influenza virus type B.

Figure 3A shows the weekly ILI and ILI+ incidence curves, illustrating their relative magnitudes over the 2024/25 winter season. Figure 3B shows the positivity rates for influenza type A and type B, highlighting their temporal patterns and relative contributions to overall influenza activity.

**Figure 3.**
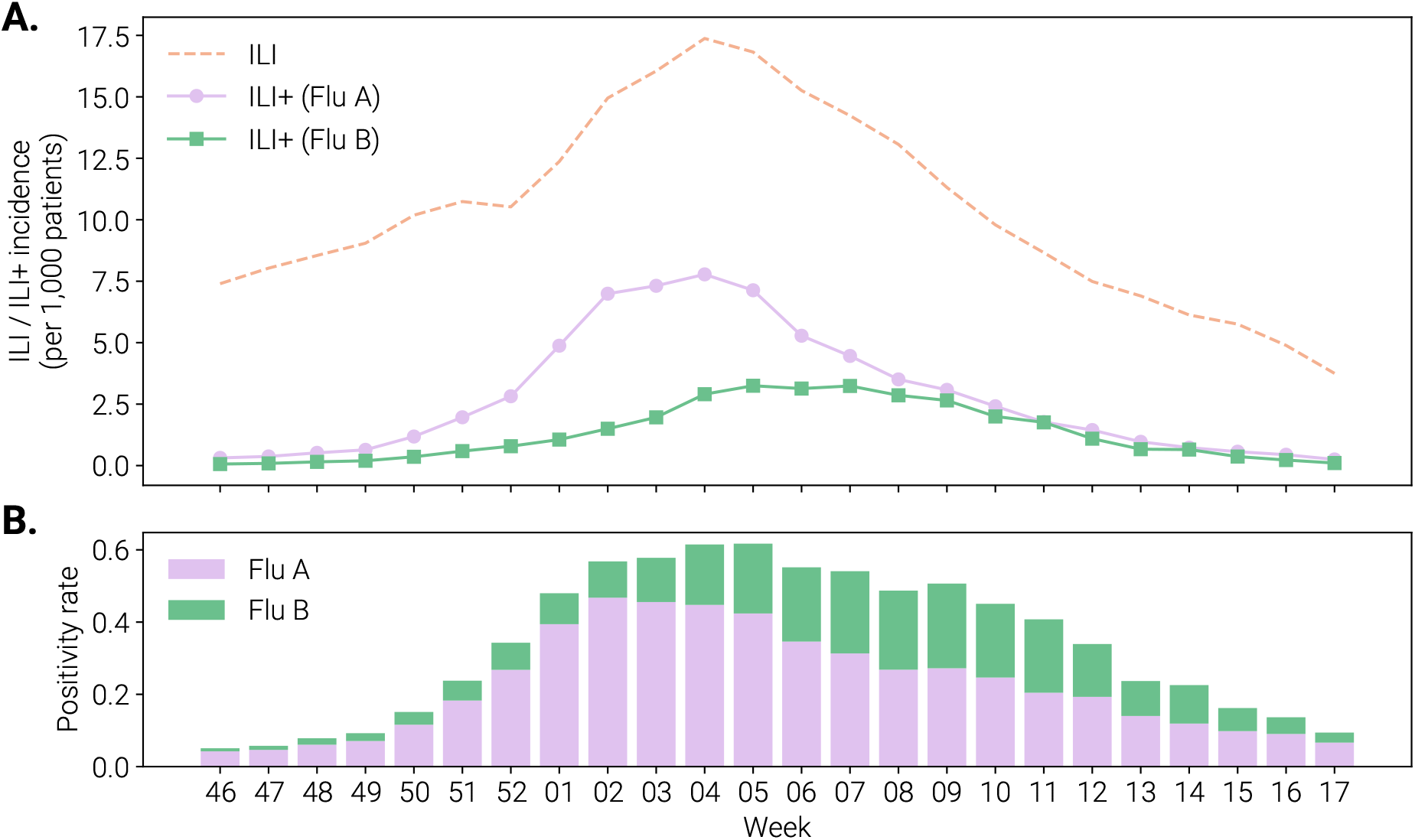
Forecasting targets for the Influcast challenge during the 2024/25 season. ILI and ILI+ signals (panel A) and positivity rate for influenza type A and type B (panel B) across the 2024/25 winter season.

### 4.3 Baseline and ensemble models

#### Baseline model

The baseline model assumes persistence of the most recent observed data, producing median forecasts equal to the last available observation in the calibration period. To generate predictive intervals, we compute one-step differences in the historical series (i.e., past changes in weekly ILI incidence) up to time *t*, denoted as *δ* = (*d*_2_, *d*_3_, …, *d*_*t*_), and symmetrize them as *δ*^*′*^ = (*δ, −δ*) to ensure centered increments. Baseline forecasts for horizon *h* are obtained by sampling *h* increments from *δ*^*′*^and summing them to the last observed value *v*_*t*_: 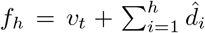. From these, 10,000 stochastic trajectories are generated to compute prediction intervals. This approach is consistent with baseline implementations adopted in other collaborative forecasting hubs [5, 6, 24, 29, 30].

#### Ensemble model

The Influcast ensemble combines individual model forecasts using a mean-across-quantiles approach, also known as the Vincent average method [23]. For each quantile of level *q*, the ensemble forecast at time *t* is computed as:

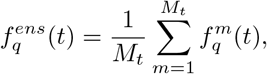

where *M*_*t*_ is the number of models submitting forecasts at time *t*. All participating models, except the baseline, are included in the ensemble construction.

We note how both the ensemble and baseline calculations follow the same approach used in the 2023/24 Influcast season [5].

### 4.4 Forecast evaluation

#### Weighted Interval Score (WIS)

The WIS is a proper scoring rule used to evaluate the accuracy and calibration of probabilistic forecasts. It provides a unified measure that summarizes how well a forecast distribution captures observed outcomes by combining two properties: *sharpness*, i.e., the narrowness of the prediction intervals, and *calibration*, i.e., the agreement between predicted and realized probabilities. Sharp and well-calibrated forecasts achieve lower WIS values. This metric has become a standard tool for comparing probabilistic forecasts in the context of infectious disease ensemble forecasting [31].

Formally, for a given forecast distribution *F* with predictive median *m*, and *K* central (1*− α*_*k*_) *×* 100% prediction intervals defined by lower and upper bounds *l*_*k*_ and *u*_*k*_, the interval score at level *α*_*k*_ is defined as:

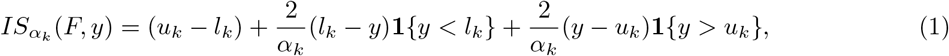

where *y* denotes the observed value and **1**{·} is an indicator function. The overall WIS is then obtained as a weighted average of the absolute error of the predictive median and the interval scores across all considered levels:

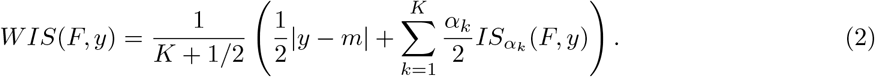

Following standard practice [31], we used *K* = 11 central prediction intervals corresponding to *α*_*k*_ = 0.02, 0.05, 0.10, 0.20, 0.30, 0.40, 0.50, 0.60, 0.70, 0.80, and 0.90.

#### Absolute Error (AE)

For each forecast instance, let *ŷ*_*i*_ denote the predicted median value and *y*_*i*_ the corresponding observed value. The AE of the median measures the magnitude of the absolute deviation between forecast and observation as:

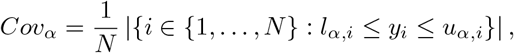

#### Prediction coverage

Prediction interval coverage is a measure of model calibration, describing how well forecast probabilities correspond to observed frequencies. It represents the proportion of observations that fall within a given prediction interval. For example, a perfectly calibrated model will have exactly 90% of observations within 90% prediction intervals. Formally, for a prediction interval of level *alpha* and a set of *N* observations *y*_*i*_, coverage is defined as:

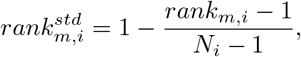

where *l*_*α,i*_ and *u*_*α,i*_ denote the lower and upper bounds of the (1 *− α*) prediction interval, respectively.

#### Relative performance

We compute the relative performance for WIS and AE using the procedure presented in Ref. [30]. For each pair of models *m* and *m*^*′*^, we compute the pairwise relative WIS or relative AE skill as:

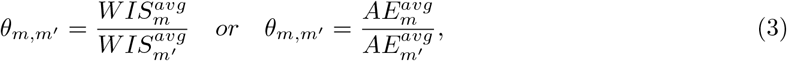

where *WIS*^*avg*^ and *AE*^*avg*^ are the average of WIS or AE of the model *m* computed using all available forecasts where both model *m* and model *m*^*′*^ participated. Then, for each model *m*, we compute the geometric mean across different *θ*_*m,m*_^*′*^ :

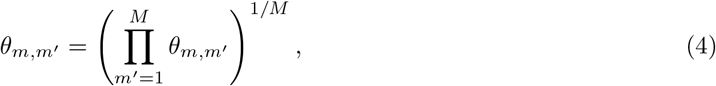

Finally, to compare the model performance with respect to the baseline model, we divide each *θ*_*m*_ by the result obtained for the baseline, named *θ*_*B*_:

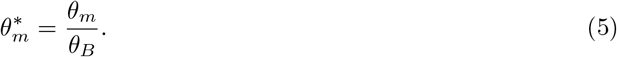

The obtained value 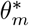 represents the relative WIS or relative AE of model *m* rescaled to the baseline model, such that the baseline has a relative performance of 1. It follows that 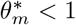 indicates that the model performs better than the baseline, and 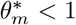 indicates that the model performs worse than the baseline.

#### Standardized rank

For each model *m* and observation *i*, the standardized rank quantifies the relative performance of the model compared to all others. It is defined as:

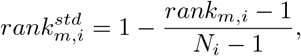

where *N*_*i*_ denotes the number of models providing forecasts for observation *i*, and rank_*m,i*_ is the rank assigned to model *m* for that observation according to the evaluation metric (either WIS or AE). The best-performing model for a given observation, corresponding to the lowest WIS (or AE), receives rank_*m,i*_ = 1 and thus a standardized score of 1, while the worst-performing model (rank_*m,i*_ = *N*_*i*_) receives a score of 0.

## Acknowledgements

S.F., E.V., M.M., C.G., P.M., D.P., M.Q., L.R., I.V., N.G. acknowledge support from the Lagrange Project of the Institute for Scientific Interchange Foundation (ISI Foundation) funded by Fondazione Cassa di Risparmio di Torino (Fondazione CRT). T.B. and V.d’A. acknowledge partial financial support from the INFN, Italy grant “LINCOLN”. M.D.D. acknowledges partial financial support from the MUR - PNC (DD n. 1511 30-09-2022) DigitAl lifelong pRevEntion (DARE) Project no. PNC0000002 and from MUR funding within the PRIN 2022 PNRR (DD n. 1214 31-07-2023) Project no. P2022A889F. R.M. and C.P. acknowledge the Cariparo Foundation, through the programme Starting Package. M.S. and C.P. acknowledge the Department of Molecular Medicine, through the programme SID from BIRD funding.

## Authors contributions

Stefania Fiandrino: Writing – review & editing, Writing – original draft, Software, Methodology, Investigation, Formal analysis. Tommaso Bertola: Writing – review & editing, Methodology, Data curation. Valeria D’Andrea: Writing – review & editing, Methodology, Data curation. Manlio De Domenico: Writing – review & editing, Methodology, Data curation. Elisa Viola: Writing – review & editing, Methodology, Data curation. Lorenzo Zino: Writing – review & editing, Methodology, Data curation. Mattia Mazzoli: Writing – review & editing, Methodology, Data curation. Alessandro Rizzo: Writing – review & editing, Methodology, Data curation. Yuhan Li: Writing – review & editing, Methodology, Data curation. Nicola Perra: Writing – review & editing, Methodology, Data curation. Marika Sartore: Writing – review & editing, Methodology, Data curation. Chiara Poletto: Writing – review & editing, Methodology, Data curation. Alberto Mateo Urdiales: Writing – review & editing, Validation, Supervision. Antonino Bella: Writing – review & editing, Validation, Supervision. Corrado Gioannini: Writing – review & editing, Software, Project administration, Conceptualization. Paolo Milano: Software. Daniela Paolotti: Writing – review & editing, Project administration, Conceptualization. Marco Quaggiotto: Visualization, Conceptualization. Luca Rossi: Writing – review & editing, Methodology, Data curation, Conceptualization. Ivan Vismara: Visualization, Software. Alessandro Vespignani: Writing – review & editing, Validation, Supervision, Project administration, Methodology, Conceptualization. Nicolò Gozzi: Writing – review & editing, Writing – original draft, Supervision, Project administration, Methodology, Investigation, Formal analysis, Conceptualization.

## Data availability

All forecasts are available at https://github.com/Predizioni-Epidemiologiche-Italia/Influcast.

