## Supplementary Information for "Insights from the second season of collaborative influenza forecasting in Italy with updated targets incorporating virological information"

### Contents

**S1 Participating models** **4**

**S2 Individual and ensemble models performance on ILI+ targets (Supplementary Analyses)** **8**

**S3 Comparison between ILI and ILI+ ensemble forecasts performance (Supplementary Analyses)** **9**

### S1 Participating models

In this section, we provide an overview of the individual models that participated in the 2024/25 Influcast forecasting season. A total of 7 teams contributed 12 models, which we grouped into three main categories according to their methodological approach: i) *mechanistic models*, which explicitly describe the biological and epidemiological processes underlying disease transmission (e.g., compartmental models); ii) *semi-mechanistic models*, which combine mechanistic assumptions with statistical or data-driven components (e.g., models with non-parametric time-varying transmission rates); and iii) *statistical models*, which do not rely on explicit transmission mechanisms but instead apply statistical or machine learning methods to identify patterns and make forecasts directly from observed data (e.g., time-series models).

Table S1 presents the list of participating models in the 2024/25 Influcast challenge along with the number of rounds submitted for each target. We also report whether each model was included in the ensemble forecasts presented in the main text. Only models that submitted all three targets were included in the main-text ensembles to ensure a fair comparison across targets. Models marked with an asterisk contributed forecasts for ILI+ targets retrospectively. Specifically, *Mechanistic-3* contributed one retrospective round, *Mechanistic-4* six rounds, *Mechanistic-5* twenty rounds, and *Statistical-1* nine rounds. The last three rounds were excluded from the evaluations because not all forecasting horizons could be evaluated against the observed data due to the end of the official surveillance.

| **Model Tag** | **Model Name** | **ILI n. Rounds** | **ILI+(Flu A) n. Rounds** | **ILI+(Flu B) n. Rounds** | **Included** |
|---|---|---|---|---|---|
| *Mechanistic-1* | *FluABCaster* | 23 | 20 | 20 | Yes |
| *Mechanistic-2* | *FluBcast* | 24 | 21 | 21 | Yes |
| *Mechanistic-3* | *GLEAM* | 24 | 20* | 20* | Yes |
| *Mechanistic-4* | *SEEIIRS_MCMC* | 23 | 21* | 21* | Yes |
| *Mechanistic-5* | *mobnetSI2R* | 24 | 21* | 21* | Yes |
| *Mechanistic-6* | *SEIRaugment* | 21 | / | / | No |
| *Mechanistic-7* | *SEIR* | 21 | / | / | No |
| *Mechanistic-8* | *metaFlu* | 21 | / | / | No |
| *Semi-mechanistic-1* | *DeepRE* | 4 | / | / | No |
| *Semi-mechanistic-2* | *REST_HE* | 21 | / | / | No |
| *Statistical-1* | *ARIMA_QMUL* | 20 | 21* | 21* | Yes |
| *Statistical-2* | *IPSICast* | 21 | / | / | No |

Table S1: **Summary of participating models in the 2024/25 Influcast challenge**. Table reports the model tag used to identify each model according to their type in the main text, the corresponding model name on the Influcast platform, the number of contributed rounds for each target, and whether the model was included or not in the main text analyses. The asterisk indicates that a model contributed some forecasting rounds retrospectively.

Below we provide a short description for each of the contributing models.

***Mechanistic-1*** (FluABCaster). Stochastic, age-structured compartmental model considering a SEIR compartmentalization setup. The population is stratified into 10 age groups ($0-9$, $10-19$, $20-24$, $25-29$, $30-39$, ..., $70-79$, $80+$) and contacts between different age groups are described by a synthetic contact matrix from Ref. [1]. The model also considers seasonality terms modulating the force of infection. The model is implemented in discrete time (the simulation step is 1 day) and the number of individuals transitioning among compartments is simulated as chain binomial processes. Model calibration is conducted using an Approximate Bayesian Computation technique [2]. Free parameters include transmissibility rate, reporting fraction, and initial immunity.

***Mechanistic-2*** (FluBcast). Stochastic, age-structured compartmental model with a standard SEIR framework and an additional compartment $S_b$ representing susceptible individuals who engage in risk-averse behaviors. These individuals experience a reduced force of infection, modulated by a parameter $r_b < 1$ that reflects the effectiveness of preventive measures (i.e., mask-wearing, reducing the number of contacts, ...). The transition from $S$ (healthy and susceptible individuals) to $S_b$ occurs at a rate influenced by the number of infectious contacts. Conversely, susceptible-behavioral individuals may revert to the susceptible compartment and relax their behavior at a rate $\mu_b$, which depends on the overall prevalence of susceptible and recovered individuals in the population. The model employs a stochastic approach, with compartment transitions modeled using chain binomial processes. The population is stratified into 10 age groups: 0–9, 10–19, 20–24, 25–29, 30–39, 40–49, 50–59, 60–69, 70–79, 80+, and contacts between age groups are described by a synthetic contact matrix [1].

***Mechanistic-3*** (GLEAM). Stochastic, age-structured compartmental model based on a metapopulation approach for simulating the spatiotemporal evolution of the spreading of infectious diseases on a global scale [3, 4]. It uses datasets derived from real-world data about worldwide population density and demographic structure, flight networks and passenger distribution, daily commuting flows, and age-structured contact patterns. The model considers 16 age groups and related contact matrices corresponding to four different settings. The epidemic dynamics within each subpopulation are defined by describing the compartmental structure of a given disease along with the corresponding transition rates and other relevant parameters.

***Mechanistic-4*** (SEEIIRS_MCMC). A deterministic SEEIIR compartmental model, an extension of the standard SEIR model in which the exposed (E) and infectious (I) compartments are duplicated to allow for an Erlang distribution of the residency times in the exposed and infectious compartments, more realistic than the exponential distribution. To capture temporal variations in transmission, the model incorporates seasonality by modeling a time-varying transmissibility according to [4]. Additionally,the model accounts for the waning of immunity, and assumed that detected cases are only a fraction of cases, determined by a detection probability p. The average durations of the latent period (2 days), the infectious period (3 days), and the immunity period (1 year) were pre-selected from a range of values available in the literature and then selected based on the model's ability to reproduce historical ILI incidence in Italy. The remaining parameters, including the initial number of individuals in each compartment, transmissibility, seasonality-related parameters, and detection probability, were estimated using a Markov Chain Monte Carlo (MCMC) approach via the Metropolis-Hastings algorithm. Non-informative uniform priors for all fitted parameters were employed. On a weekly basis, the model has been fitted to ILI incidence. The resulting posterior distributions were then used to generate trend predictions for the following four weeks, presented as prediction intervals. This same procedure was applied independently to ILI+(Flu A) and ILI+(Flu B) targets.

***Mechanistic-5*** (mobnetSI2R). Region-based SIR metapopulation model that incorporates mobility fluxes of individuals between regions in the force of infection [5]. The model considers two strains of pathogens contributing to the overall incidence. Mobility data between regions is estimated using a radiation model calibrated on the population data from the last available census. The assumptions on mobility allow for obtaining reasonable estimates even for regions not provide updated data. The model consists of a system of ODEs solved using a Runge-Kutta 4th order algorithm with a daily time step and is calibrated using a particle swarm optimisation algorithm by considering in the fitness the overall incidence, the incidence in each region, and the ratios of the two strains at the national level. Uncertainty quantification is performed by repeating the calibration 1000 times and gathering statistics on the obtained results.

***Mechanistic-6*** (SEIRaugment). An age-structured SEIR model augmented with outputs from a statistical ARIMA model (statistical-1). Four-week-ahead forecasts from the ARIMA model for the current week are used as additional calibration targets alongside historical data. The model is calibrated using a simple Approximate Bayesian Computation (ABC) method where the top 1% are accepted, and the weighted mean absolute percentage error (wMAPE) is used as the target metric for calibration.

***Mechanistic-7*** (SEIR). Age-structured SEIR model calibrated using a simple Approximate Bayesian Computation (ABC) method where the top 1% (out of $200,000$ simulations) are accepted. We used the weighted mean absolute percentage error (wMAPE) as the target metric for calibration.

***Mechanistic-8*** (metaFlu). Province-based metapopulation model built starting from the model used for COVID-19 in Ref. [6]. A distribution for the model parameters related to the disease progression has been obtained by calibrating the model to past year influenza seasons and adjusted using available data on the 2023/24 influenza season. To generate the predictions, we simulated 1000 realizations of the dynamical system, each one with an independent realization of the model parameters, sampled from the distributions obtained in the calibration process.

***Semi-mechanistic-1*** (DeepRE). Deep Renewal Equation is a model coupling Renewal Theory and Deep Learning. The intuition is that renewal equations in the context of epidemic forecasting are characterized by 3 components: the reproductive factor, the historical series of infected, and the generation time distribution. Putting aside the historical data, which can be assumed to be known, we approximate the other two components of the equation through deep neural networks, allowing for data-driven approximation of these components. The model can be used in two ways: on the one hand, we can predict the epidemic trend, namely the number of infected in time; while on the other hand, the approximated reproductive factor and the generation time distribution can be used to better understand the epidemic itself.

***Semi-mechanistic-2*** (REST_HE). The model is based on the application of the Renewal equation and assumes a distribution of the generation time that is intermediate between those of SARS-CoV-2 and seasonal flu. The final projections are the ensemble of projections based on two different assumptions: i) the reproduction number remains constant at a fixed value (corresponding to the last available estimate) throughout the projection horizon; ii) the reproduction number decreases over time from the last available estimate, with the same trend observed in data from 2003/04 to 2022/23, excluding outlier seasons due to the H1N1 and COVID-19 pandemics (2009/10, 2020/21 and 2021/22). The model takes into account both the stochastic variability of the incidence and the uncertainty in the estimate of the reproduction number.

***Statistical-1*** (ARIMA_QMUL). Autoregressive integrated moving average (ARIMA) model trained by using data from the previous (2022/23) and current (2023/24) influenza season.

***Statistical-2*** (IPSICast). Linear autoregressive exogenous model in which the forecasts are based on past official data and an exogenous variable. The traditional surveillance data comes from the official national institute. The exogenous variable comes from the information retrieved from Influweb, a web-based participatory surveillance platform that has been monitoring ILI incidence in Italy since 2008 as part of the Influenzanet network [7, 8]. Influweb's participatory system offers the advantage of providing real-time data, a feature not present with traditional surveillance, which only offers data for the preceding week. Leveraging this real-time information, this model incorporates the instantaneous ILI signal from Influweb to forecast the upcoming four weeks. The approach involves generating predictions for ILI by integrating the ILI incidence official data from the three preceding weeks and the current week's signal from Influweb, averaged with the two preceding weeks, into a linear autoregressive exogenous model. The regression coefficients of the autoregressive model are estimated separately for the different time horizons, using a least squares regression.

#### S2 Individual and ensemble models performance on ILI+ targets (Supplementary Analyses)

Figure S1 reports the standardized ranks of the absolute error (AE) of the median across different forecast horizons and rounds for ILI+ targets. For ILI+(Flu A), *Mechanistic-1*, *Mechanistic-2*, and *Mechanistic-3* achieved the highest median AE standardized ranks, all sharing the same median value, followed by the *Ensemble*. For ILI+(Flu B), the *Ensemble* achieved the highest median AE standardized rank, followed by *Mechanistic-3*, *Mechanistic-1*, and *Mechanistic-2*. According to the AE standardized rank, the *Ensemble* ranked in the top half of the distribution 78% and 88% of rounds for ILI+(Flu A) and for ILI+(Flu B), respectively, indicating higher performance stability compared to any other individual model. Similar to the main text findings, the *Ensemble* also showed lower variability in performance across rounds compared to other top-performing models.

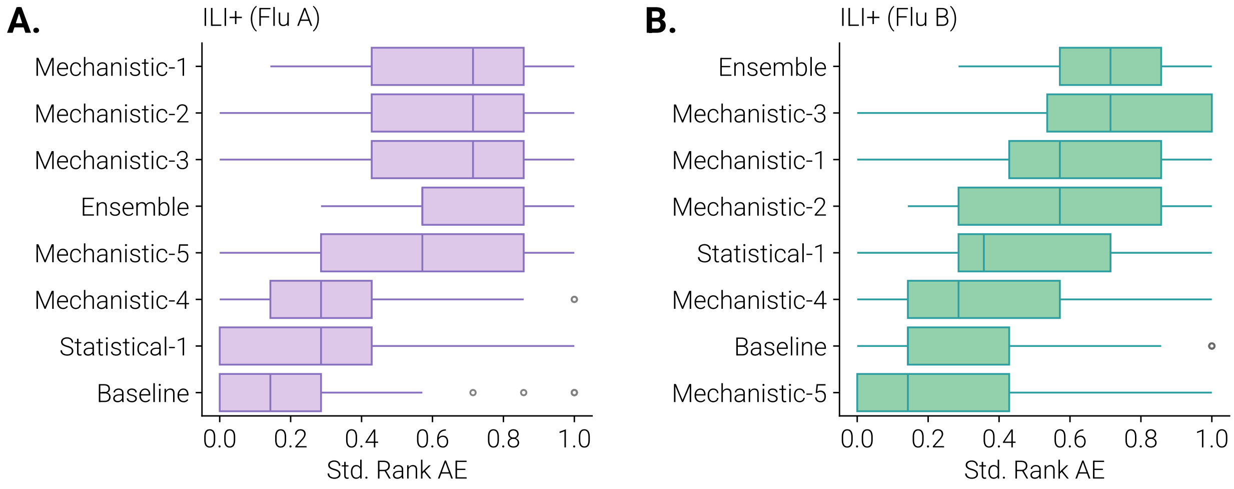


Figure S1: **Standardized AE rank of 2024/2025 Influcast forecasting models for ILI+(Flu A) and ILI+(Flu B)**. Plot shows the boxplots of standardized AE ranks of different models computed combining different horizons and forecasting rounds for ILI+(Flu A) (panel A) and ILI+(Flu B) (panel B). The boxplot boundaries represent the interquartile range (IQR) spanning between the first quartile (Q1) and third quartile (Q3), while the line inside each box indicates the median. The whiskers extend to the furthest data point within 1.5 times the IQR from Q1 and Q3. Outliers are shown as individual points.

### S3 Comparison between ILI and ILI+ ensemble forecasts performance (Supplementary Analyses)

Figure S2 displays the one- to four-week-ahead forecasts for the three targets, ILI+(Flu A), ILI+(Flu B), and ILI, showing the median together with the 50% and 90% predictive intervals (panels A, B, and C, respectively). Across all horizons, the ensemble generally tracks the temporal patterns of the ILI+ targets well. In particular, for both Flu A and Flu B (panels A and B), the ensemble captures the early season rise and the timing of the peak reasonably accurately, although performance naturally degrades at longer horizons. At horizons 3 and 4, we observe wider predictive intervals and a tendency to underpredict during the initial growth phase, reflecting greater uncertainty. A similar pattern is evident for the ILI forecasts (panel C). In this case, however, the ensemble already struggles to capture the early-season increase at the short horizons, with the underestimation becoming even more pronounced at horizons 3 and 4. After the peak, forecast performance improves, with narrower predictive intervals and better alignment with the observed values. Overall, the figure illustrates that the ensemble reliably reproduces short-term dynamics for all targets, with the expected decline in accuracy and widening uncertainty at longer horizons, and with generally better alignment for ILI+ targets than for ILI.

Figures S3 and S4 summarize the performance of the individual models for the ILI and ILI+ targets, using WIS and AE aggregated across all forecasting rounds and horizons. Overall, all models show better median WIS and AE for the ILI+(Flu A) target than for ILI. The largest and statistically significant improvements (Wilcoxon signed-rank test, 1% level) are observed for *Mechanistic-3*, *Mechanistic-4*, and *Mechanistic-5*. *Mechanistic-3* shows a reduction in median WIS from ILI (1.13) to ILI+(Flu A) (0.38), and a decrease in AE from 1.08 to 0.31. *Mechanistic-4* similarly improves from 1.25 to 0.86 in WIS and from 1.08 to 0.72 in AE, and *Mechanistic-5* from 0.86 to 0.53 in WIS and from 0.72 to 0.55 in AE. For all other models, differences between ILI and ILI+(Flu A) are not statistically significant. For the ILI+(Flu B) target, three models, *Mechanistic-3*, *Mechanistic-4*, and *Statistical-1*, show improved median WIS and AE relative to ILI. Of these, only *Mechanistic-3* and *Mechanistic-4* exhibit statistically significant improvements at the 1% level (Wilcoxon signed-rank test), except for AE for *Mechanistic-4*, which is significant at the 10% level. Table S2 reports the average 90% coverage across forecasting rounds for the ILI, ILI+(Flu A), and ILI+(Flu B) targets at the individual model level. Overall, we observe improved calibration for both ILI+ targets compared with ILI. Three models, *Mechanistic-3*, *Mechanistic-4*, and *Statistical-1*, achieve higher coverage for both ILI+(Flu A) and ILI+(Flu B) than for ILI. Two additional models, *Mechanistic-2* and *Mechanistic-5*, show higher coverage for ILI+(Flu A) but not for ILI+(Flu B). The only exception is *Mechanistic-1*, which exhibits higher coverage for ILI than for either ILI+ target.

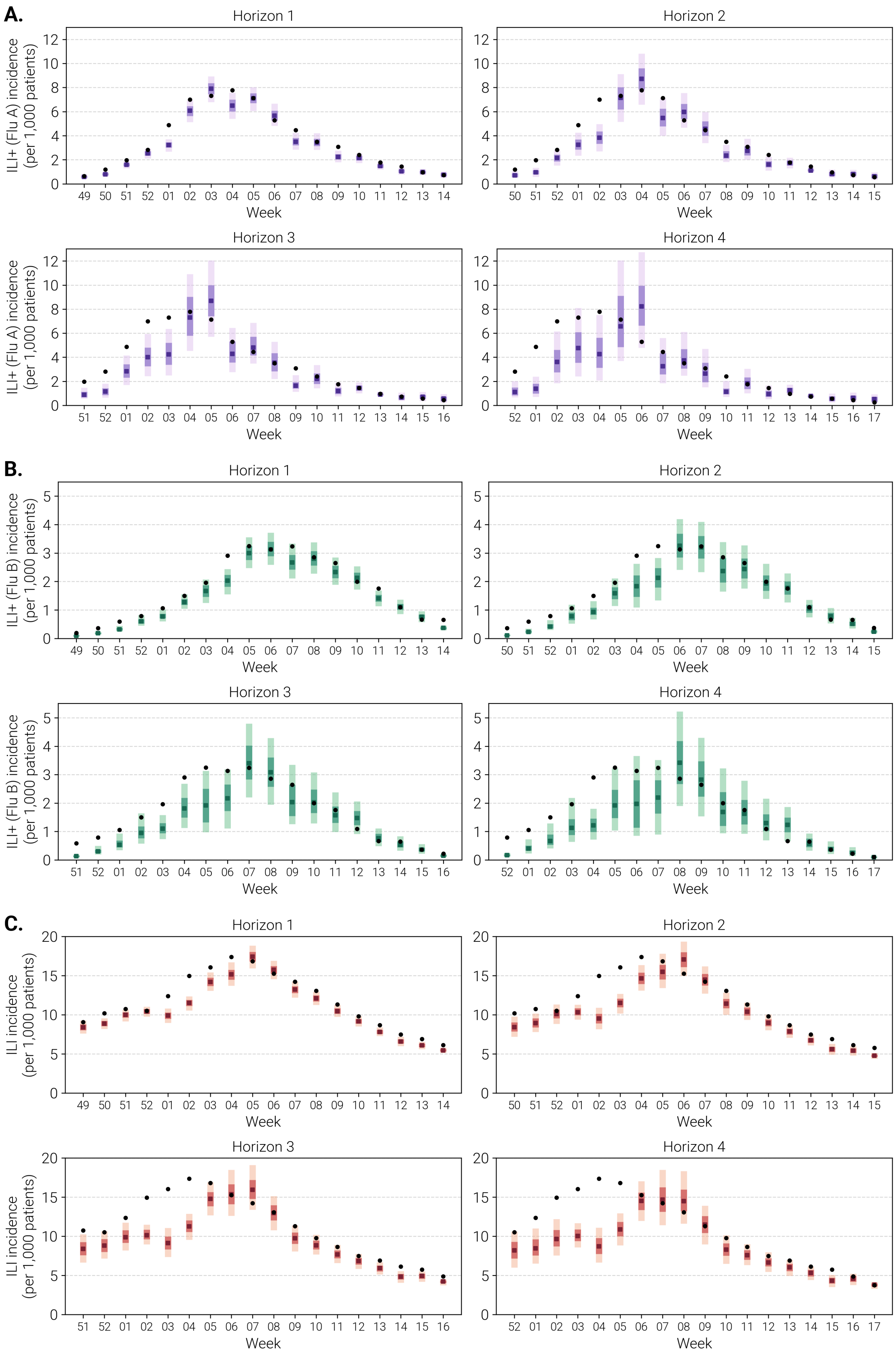


Figure S2: **Influcast ensemble one- to four-week-ahead forecast.** The plot shows the one- to four-week-ahead forecasts, presenting the median, 50%, and 90% prediction intervals for ILI+(FluA) (panel **A**), ILI+(FluB) (panel **B**), and ILI (**C**).

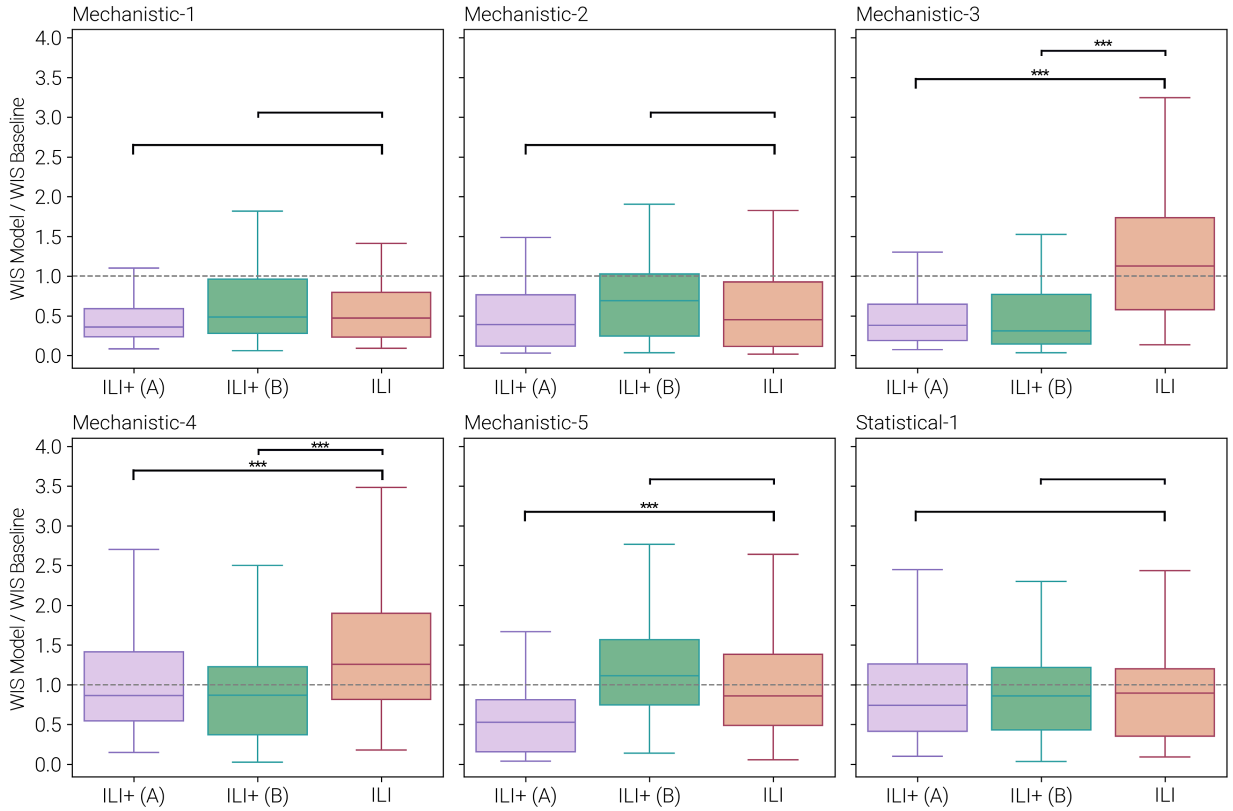


Figure S3: **Individual models forecasting performance on ILI and ILI+, WIS**. WIS ratio between the individual model and the Baseline model computed by aggregating across forecasting rounds and horizons. Boxplots show the distribution of WIS values. The boxplot boundaries represent the interquartile range (IQR) between the first quartile (Q1) and third quartile (Q3), while the line inside each box indicates the median. The whiskers extend to the furthest data point within 1.5 times the IQR from Q1 and Q3. In all the panels, we report the statistical significance of the Wilcoxon signed-rank test comparing ILI versus ILI+(Flu A) and ILI+(Flu B) as follows: $^{***}$: $p_{val} < 0.01$, $^{**}$: $0.01 < p_{val} < 0.05$, $^{*}$: $0.05 < p_{val} < 0.1$, and blank otherwise.

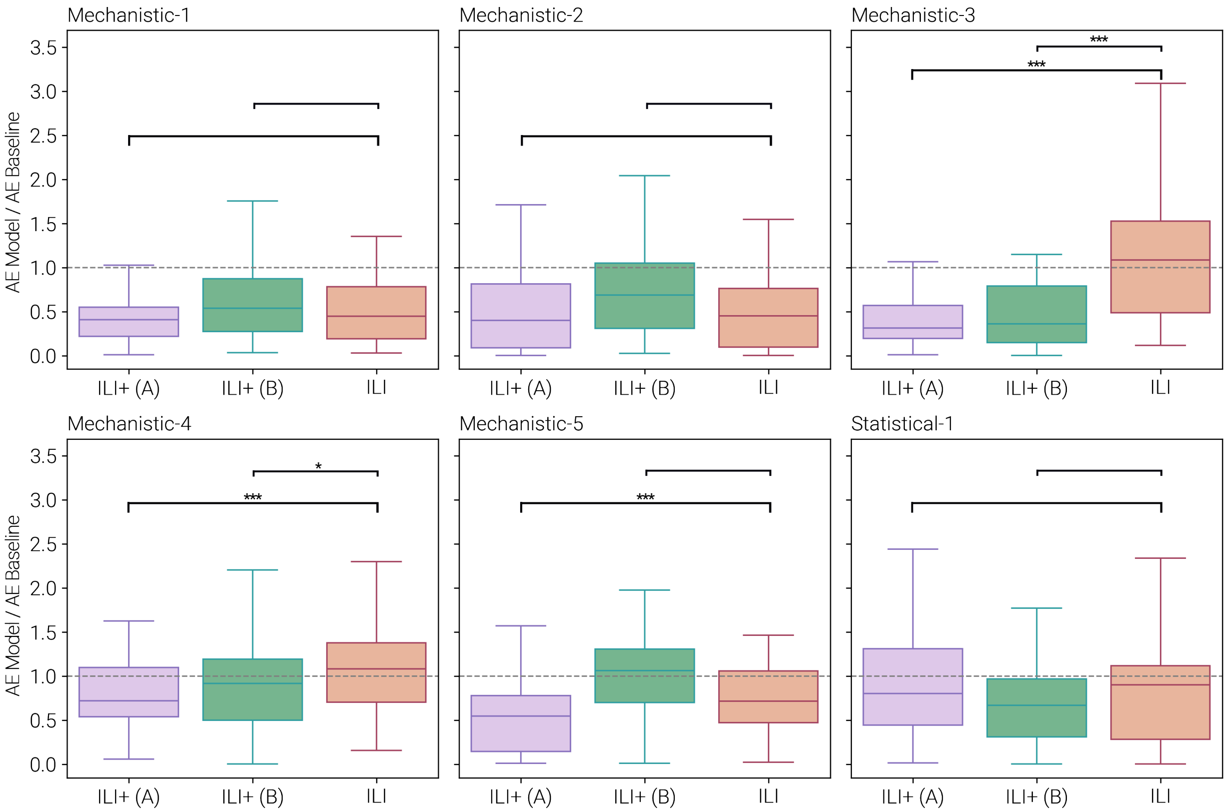


Figure S4: **Individual models forecasting performance on ILI and ILI+, AE**. AE ratio between the individual model and the Baseline model computed by aggregating across forecasting rounds and horizons. Boxplots show the distribution of AE values. The boxplot boundaries represent the interquartile range (IQR) between the first quartile (Q1) and third quartile (Q3), while the line inside each box indicates the median. The whiskers extend to the furthest data point within 1.5 times the IQR from Q1 and Q3. In all the panels, we report the statistical significance of the Wilcoxon signed-rank test comparing ILI versus ILI+(Flu A) and ILI+(Flu B) as follows: $^{***}$: $p_{val} < 0.01$, $^{**}$: $0.01 < p_{val} < 0.05$, $^{*}$: $0.05 < p_{val} < 0.1$, and blank otherwise.

| **Model Tag** | **90% Coverage** | | |
|---|---|---|---|
| | ILI+(Flu A) | ILI+(Flu B) | ILI |
| *Mechanistic-1* | 0.60 | 0.62 | 0.74 |
| *Mechanistic-2* | 0.53 | 0.43 | 0.51 |
| *Mechanistic-3* | 0.58 | 0.74 | 0.22 |
| *Mechanistic-4* | 0.24 | 0.39 | 0.16 |
| *Mechanistic-5* | 0.47 | 0.14 | 0.36 |
| *Statistical-1* | 0.49 | 0.44 | 0.42 |

Table S2: **Individual models forecasting performance on ILI and ILI+, 90% coverage**. The Table reports the 90% coverage values averaged across forecasting rounds, for three forecasting targets: ILI+(Flu A), ILI+(Flu B), and ILI.

We conducted a first sensitivity analysis using the ensemble restricted to models that submitted forecasts for all three targets (ILI, ILI+(Flu A), and ILI+(Flu B)), without requiring the comparison to rounds common to all targets. Under this setup, we include three additional ILI forecast rounds, weeks 2024-45 to 2024-47, for which ILI+(Flu A) and ILI+(Flu B) submissions were unavailable due to missing data. Figures S5A and S5B show the distributions of WIS and AE ratios between the Ensemble and the Baseline models, aggregated across horizons and stratified by horizon, for the ensemble across the three targets. The findings are consistent with those from the main analysis. Even with the additional rounds included, the ensemble's aggregated performance (left panels) shows a clear pattern: the median ratio of the WIS between the Ensemble and the Baseline models for the ILI target (0.74) remains higher than for ILI+(Flu A) (0.36) and ILI+(Flu B) (0.47), indicating improved forecast accuracy for both ILI+ targets. Similar patterns emerge when performance is examined by forecast horizon, with differences again most pronounced for the ILI+(Flu A) target. Because the additional ILI rounds break the pairing between targets, we evaluate differences between ILI and the two ILI+ targets using the Mann–Whitney U test (Wilcoxon rank-sum test). For aggregated WIS and AE, the improvements for both ILI+(Flu A) and ILI+(Flu B) relative to ILI are statistically significant at the 1% level. When disaggregating by forecast horizon, the strength of the differences varies across targets. For ILI+(Flu A), relative performance remains significantly better than ILI at the 1% level for horizons 1, 2, and 3, but differences at horizon 4 are not statistically significant for either WIS or AE. For ILI+(Flu B), differences are weaker: significance is observed only at the 10% level for horizons 2 and 3, while horizons 1 and 4 show no significant differences. Figure S5C shows the corresponding 90% coverage values by forecasting round. Also in this case, even considering three additional rounds for ILI, the dashed horizontal lines representing the average coverage across all rounds show, on average, improved calibration of the ensemble for the ILI+ targets, with value 0.54 for ILI, compared to 0.60 for ILI+(Flu A), and 0.64 for ILI+(Flu B).

We conducted a second sensitivity analysis using the official ensemble for ILI incidence, which includes

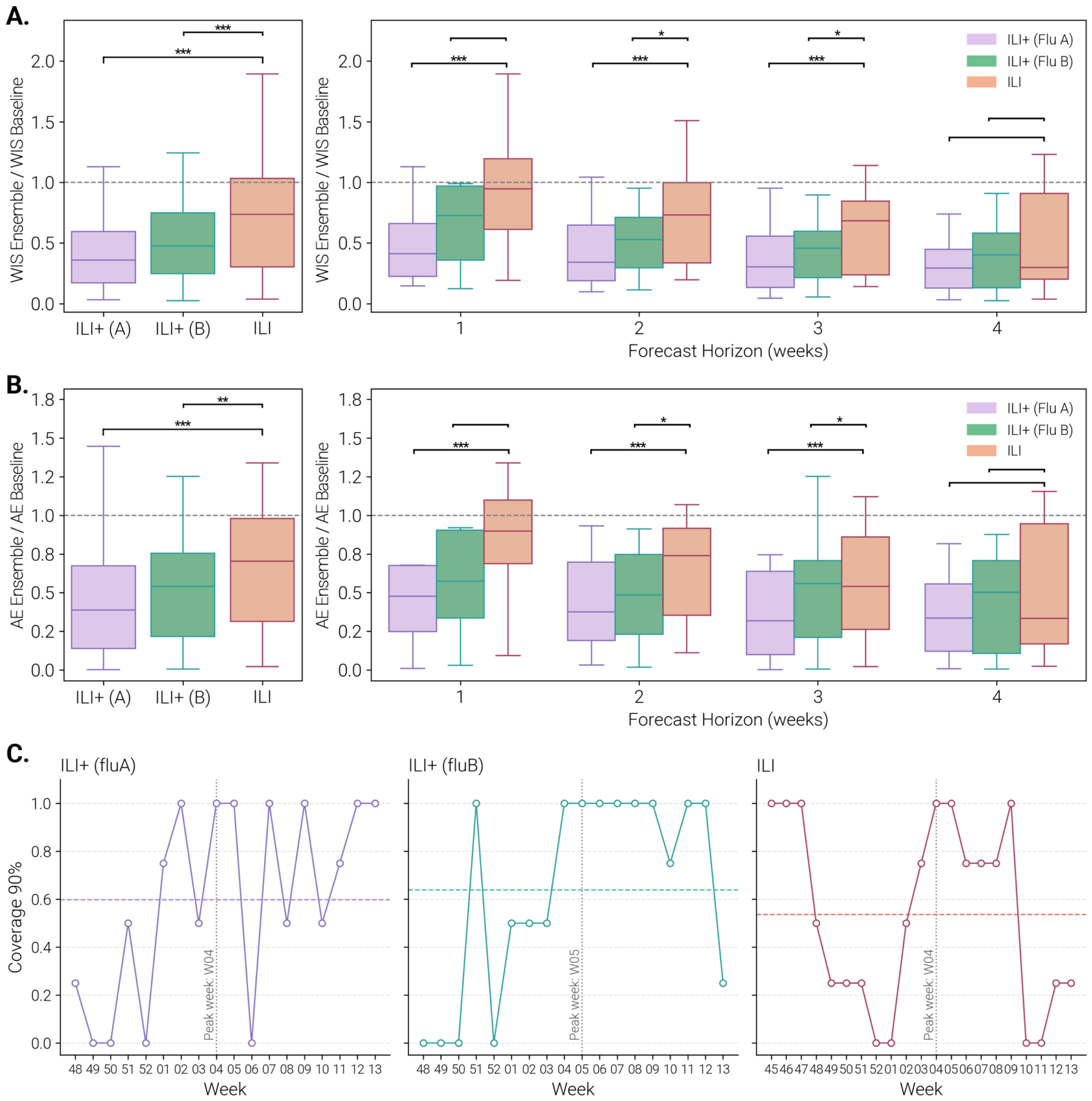


Figure S5: **Sensitivity analysis on number of included rounds: ensemble forecasting performance on ILI and ILI+.** WIS ratio **(A)** and AE ratio **(B)** between the Ensemble and the Baseline models computed aggregating across forecasting rounds and horizons (left panels) and disaggregated by horizons (right panels). Boxplots show the distribution of WIS/AE values. The boxplot boundaries represent the interquartile range (IQR) between the first quartile (Q1) and third quartile (Q3), while the line inside each box indicates the median. The whiskers extend to the furthest data point within 1.5 times the IQR from Q1 and Q3. In all the panels, we report the statistical significance of the Mann–Whitney U test (Wilcoxon rank-sum test) comparing ILI versus ILI+(Flu A) and ILI+(Flu B) as follows: $^{***}$: $p_{val} < 0.01$, $^{**}$: $0.01 < p_{val} < 0.05$, $^{*}$: $0.05 < p_{val} < 0.1$, and blank otherwise. **(C)** 90% coverage values in different rounds for ILI, ILI+(Flu A), and ILI+(Flu B). Dashed horizontal lines represent the average value across forecasting rounds, while the grey vertical dashed line indicates the week of the peak of each target.

all submitted models regardless of whether they provided forecasts for all targets. This affects only the ILI target, as it allows additional models that submitted forecasts exclusively for ILI; the ILI+(Flu A) and ILI+(Flu B) ensembles remain unchanged. In this configuration, twelve models contribute to the ILI forecasts. Figures S6A and S6B present the distributions of WIS and AE ratio between the Ensemble and the Baseline models, aggregated across horizons and stratified by horizon, for the three targets. Relative to both the main analysis and the first sensitivity analysis, the aggregated performance differences are even more pronounced. The median WIS ratio for ILI (0.97) remains substantially higher than for ILI+(Flu A) (0.36) and ILI+(Flu B) (0.47), and these differences are statistically significant at the 1% level. At the horizon level, the strength of these differences increases as well. For ILI+(Flu A), differences are significant at the 1% level for horizons 1, 2, and 3, and at the 5% level for horizon 4. For ILI+(Flu B), significance is observed at the 1% level for horizon 2, the 5% level for horizons 3 and 4, and is not significant for horizon 1. Figure S6C shows the 90% coverage by forecasting round. The dashed lines indicate mean coverage values across rounds, and reveal worse average calibration for the ILI target under the official ensemble (0.43) compared with ILI+(Flu A) (0.60) and ILI+(Flu B) (0.64).

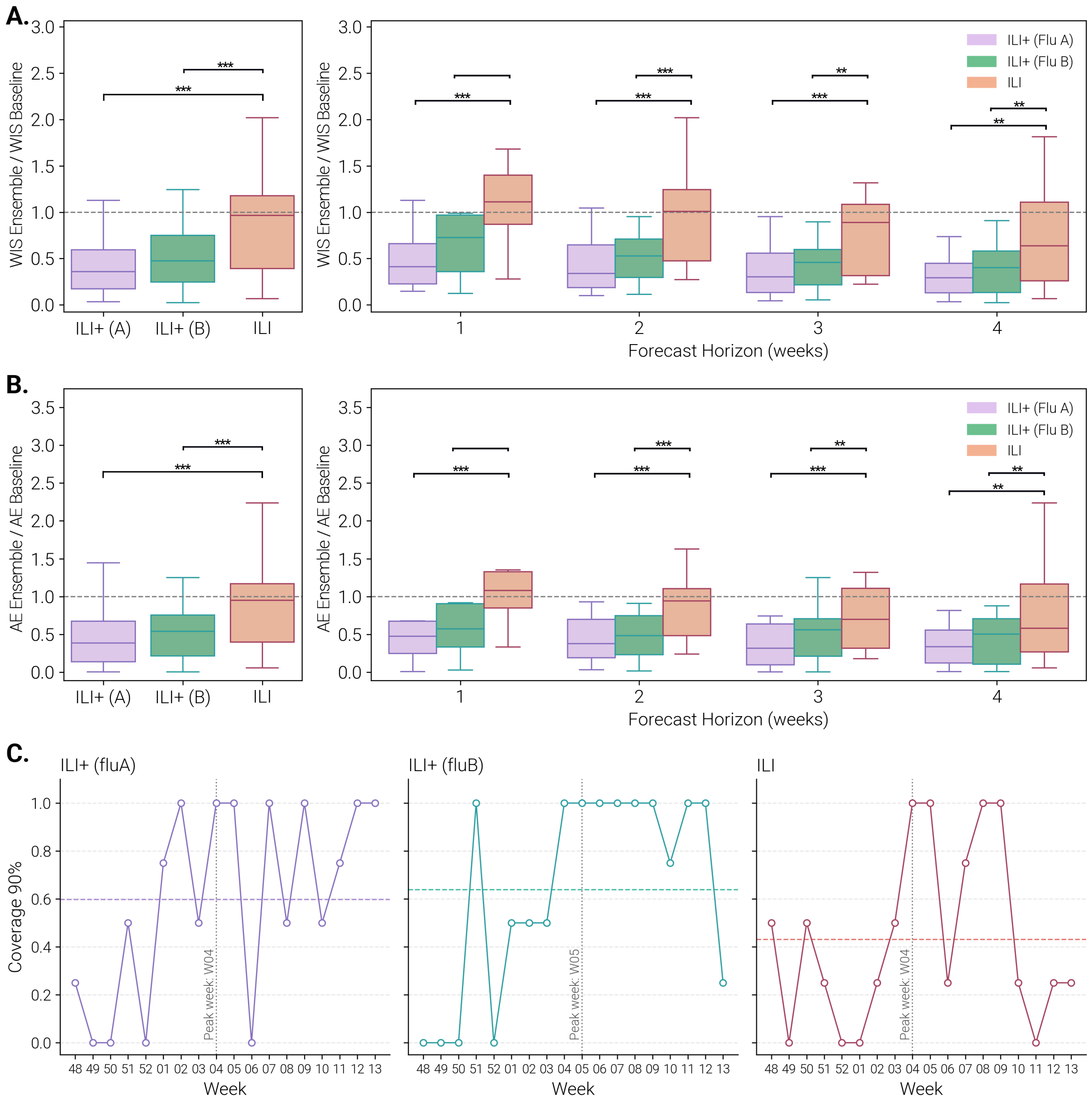


Figure S6: **Sensitivity analysis on number of included models: ensemble forecasting performance on ILI and ILI+.** WIS ratio **(A)** and AE ratio **(B)** between the Ensemble and the Baseline models computed aggregating across forecasting rounds and horizons (left panels) and disaggregated by horizons (right panels). Boxplots show the distribution of WIS/AE values. The boxplot boundaries represent the interquartile range (IQR) between the first quartile (Q1) and third quartile (Q3), while the line inside each box indicates the median. The whiskers extend to the furthest data point within 1.5 times the IQR from Q1 and Q3. In all the panels, we report the statistical significance of the Wilcoxon signed-rank test comparing ILI versus ILI+(Flu A) and ILI+(Flu B) as follows: $^{***}$: $p_{val} < 0.01$, $^{**}$: $0.01 < p_{val} < 0.05$, $^{*}$: $0.05 < p_{val} < 0.1$, and blank otherwise. **(C)** 90% coverage values in different rounds for ILI, ILI+(Flu A), and ILI+(Flu B). Dashed horizontal lines represent the average value across forecasting rounds, while the grey vertical dashed line indicates the week of the peak of each target.
